# Does directly integrating health information exchange (HIE) data with the electronic health record increase HIE use by clinicians in the emergency department?

**DOI:** 10.1101/2022.05.20.22275255

**Authors:** Heidi Hosler, Jeong Hoon Jang, Jason T. Schaffer, John Price, Titus K. Schleyer, Rebecca L. Rivera

**Author notes:** **Corresponding Author:** Heidi Hosler, MPH, Public and Population Health Informatics Pre-Doctoral Fellow, Clem McDonald Center for Biomedical Informatics, Regenstrief Institute, 1101 West 10th Street, Indianapolis, IN 46202. These authors contributed equally to this work (Titus K. Schleyer, Rebecca L. Rivera). **Author Contributions: Heidi Hosler:** Conceptualization, Formal analysis, Investigation, Methodology, Project administration, Validation, Visualization, Writing-original draft **Jeong Hoon Jang:** Formal analysis, Methodology, Validation **Jason T. Schaffer:** Funding acquisition, Resources, Writing-review & editing **John Price:** Data curation, Resources **Titus K. Schleyer:** Conceptualization, Funding acquisition, Investigation, Resources, Supervision, Validation, Visualization, Writing-review & editing **Rebecca L. Rivera:** Conceptualization; Formal analysis, Funding acquisition, Investigation, Methodology, Project administration, Supervision, Validation, Visualization, Writing-review & editing.

## Abstract

**Objective:** Develop and evaluate the effect of a Fast Healthcare Interoperability Resources (FHIR) app, *Health Dart*, integrating information from Indiana’s community health information exchange (HIE), the Indiana Network for Patient Care (INPC), directly with Cerner, an electronic health record (EHR)

**Materials and Methods:** *Health Dart* was implemented in 14 Indiana University Health emergency departments (ED) using a stepped-wedge study design. We analyzed rates of INPC use in 286,175 ED encounters between October 1, 2019 and December 31, 2020. Logistic regression was used to model the probability of INPC use given the implementation context, such as user interface (UI) enhancements and the COVID-19 pandemic.

**Results:** INPC use increased by 131% across all encounters (from 3.6% to 8.3%; p<0.001) after *Health Dart* implementation. INPC use increased by144% (from 3.6% to 8.8%; p<0.001) more than two months post-implementation. After UI enhancements, post-implementation INPC use increased 123% (from 3.5% to 7.8%) compared to 181% (from 3.6% to 10.1%; p<0.001) in post-implementation encounters that occurred before UI enhancements. During the pandemic, post-implementation INPC use increased by 135% (from 3.4% to 8.0%; p<0.001) compared to 178% (from 3.6% to 10%; p<0.001) in post-implementation encounters that occurred before the pandemic. Statistical significance was determined using 95% confidence intervals (α=0.05).

**Discussion:** Direct integration of HIE information into an EHR substantially increased frequency of HIE use, but the effect was weakened by the UI enhancements and pandemic.

**Conclusion:** HIE information integrated into EHRs in the form of dashboards can potentially make information retrieval more efficient and effective for clinicians.

## INTRODUCTION

Providing clinicians with the information they need when and in the format they need it is a significant research challenge in clinical informatics.^1–4^ Information needed to care for patients is typically still fragmented across too many different systems and not easy to collate, organize and review.^1,5,6^ This problem is aggravated in the context of increasing interoperability. Not only are clinicians tasked with thoroughly reviewing patient cases in their own electronic health record (EHR) - they are expected to do the same for patient records generated outside of their organization available through health information exchange (HIE).

HIE provides many benefits for healthcare processes and outcomes, which is why it is strongly supported by the Office of the National Coordinator and multiple other stakeholders.^7–14^ Among these benefits are fewer duplicated procedures, reduced imaging, lower costs, and improved patient safety.^7,15^ Despite these benefits, the majority of hospitals do not engage in meaningful integration of shared health data in the EHR beyond the continuity of care record.^16,17^

Clinicians ideally need *all relevant* information about a specific patient in *one* place.^18^ Missing (or inaccessible) information can have detrimental consequences for care.^19^ Community health information exchanges typically provide broad, comprehensive coverage of patient information from regional healthcare organizations, regardless of which EHR they use.^20^

Vendor-mediated HIEs, on the other hand, such as *Care Everywhere* in *Epic* and *Commonwell* in *Cerner*, and collaboratives such as *Carequality*, are often more constrained in their scale and scope of information coverage – making it sometimes difficult or impossible for clinicians to access all relevant information about a patient.^20^

Clinicians typically access information in the HIE through a separate application or portal because most HIEs are not directly integrated with EHRs. However, the resulting multiple logins, workflow interruptions, and poor presentation often impede effective and efficient information retrieval, resulting in a low level of use, especially in emergency medicine.^21–25^ Vendor-mediated HIEs often preferentially integrate external information in the EHR only when it comes from their customers, leaving information from other EHRs more difficult to access.^20^ Early evidence shows that integrating HIE information directly into the EHR can substantially increase the use of information from the HIE.^25,26^

Retrieving patient information through most HIE portals is subject to an additional limitation: Clinicians must typically browse through information organized by *type*, such as medications, labs, orders or physician notes, and *time*, because like EHRs, HIEs do not offer problem-oriented views.^27^ Problem-oriented views have been shown to improve data retrieval workflows, allowing providers to complete EHR tasks more efficiently, with fewer errors and cognitive task load, and greater user satisfaction.^28–31^

These limitations have two major consequences. First, the lack of widescale interoperability and barriers to information access imposes an exhausting litany of clerical tasks on clinicians which contributes to burnout and waste.^32,33^ Second, clinicians routinely forgo searching for and retrieving additional clinical data about patients,^34^ which contributes to waste and adverse patient outcomes.

The goal of this study, therefore, was two-fold: (1) develop a Fast Healthcare Interoperability Resources (FHIR) app, *Health Dart*, that integrates information from Indiana’s community HIE, the Indiana Network for Patient Care (INPC), directly with an EHR in the form of a problem-oriented dashboard; (2) determine how the implementation of *Health Dart* affected INPC use among clinicians at 14 Indiana University (IU) Health emergency departments (EDs) using a stepped-wedge trial design, allowing us to causally relate *Health Dart* with changes in INPC use.

## MATERIALS AND METHODS

### *Health Dart* development and pilot implementation

We developed the *Health Dart* application (app) (**Figure 1**) based on an earlier version focused on chest pain.^25^ *Health Dart* uses the FHIR standard to integrate highly relevant information from the INPC directly into *Cerner*, the EHR used at Indiana University (IU) Health and its EDs. Prior to the implementation of *Health Dart*, IU Health clinicians’ only means of accessing the INPC was via a web-based tool called *CareWeb*.^35^ After navigating to *CareWeb* from *Cerner*\*, clinicians had to browse or search through the patient’s health records to find relevant information. *Health Dart*, on the other hand, retrieves the information from the INPC that is most relevant to seven chief complaints (chest pain, abdominal pain, weakness/dizziness/headache, back/flank pain, pregnancy, arrhythmia, and dyspnea) and integrates it into *Cerner* in the form of a chief complaint-focused dashboard. A group of ED physicians, led by JS, identified the information to be displayed for each chief complaint. The supported chief complaints are the reason for approximately 40% of all encounters across the IU Health ED system and common in other EDs.^36^

**Figure 1.**
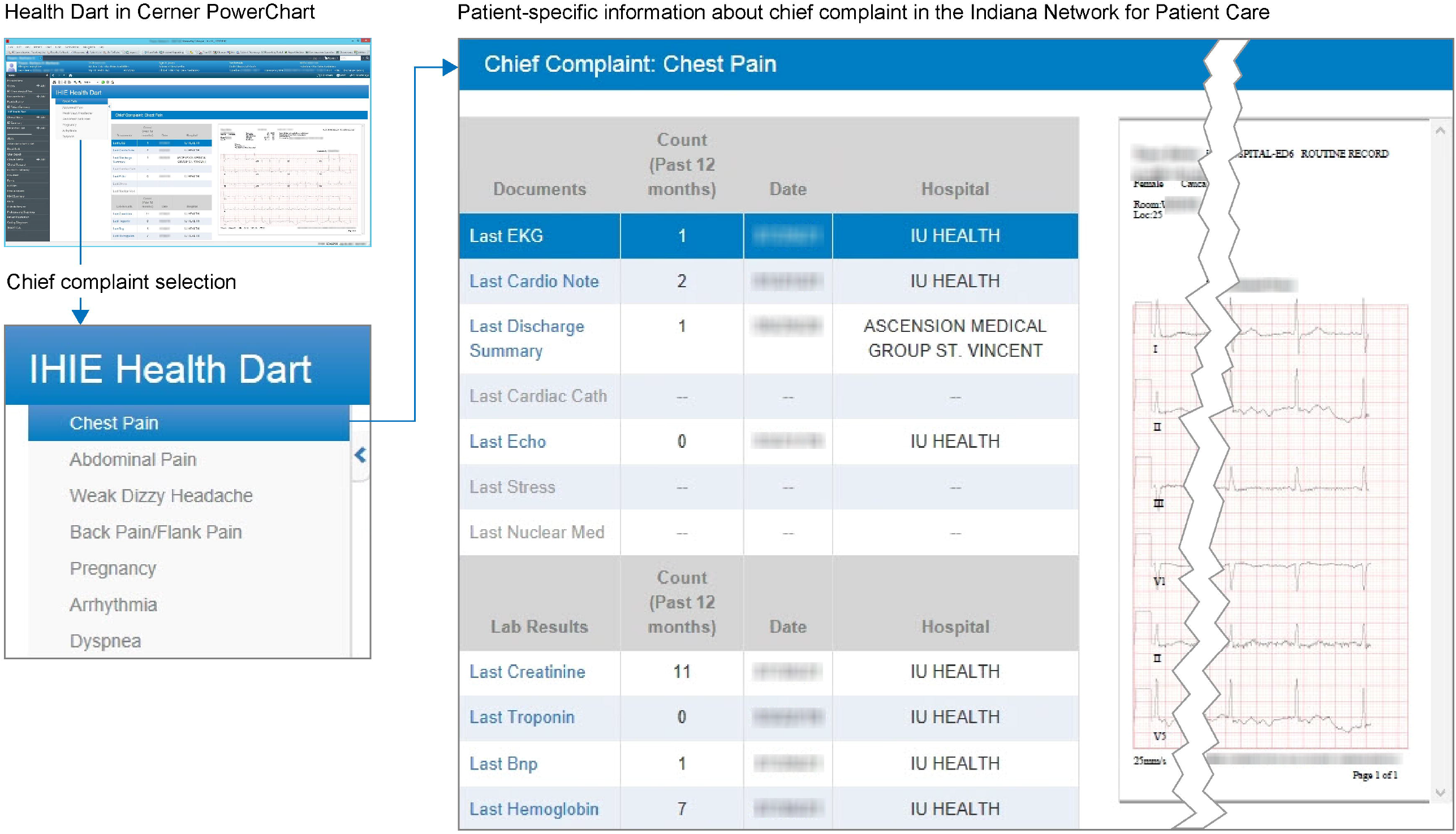
*Health Dart* screenshot displaying patient data for the chief complaint of chest pain

Clinicians access *Health Dart* directly within *Cerner PowerChart* with one click, allowing them to launch the app without navigating to *CareWeb*, and possibly having to re-enter user credentials and search for/select a patient. *Health Dart* is an *additional* way to access information in the INPC. The traditional access to *CareWeb* is still available if clinicians need to review the full patient record in the INPC application. *Health Dart* was implemented at a central pilot site, IU Health’s Methodist ED, in January 2018 before its launch across the whole IU Health system.

### Study Design and Setting

After pilot implementation, *Health Dart* was implemented in 14 IU Health EDs with 166 clinicians using a stepped-wedge, cluster non-randomized controlled study design. As an alternative to randomized controlled trials, which are often not practical for site-based studies of healthcare delivery interventions, this scientifically validated, pragmatic study design enables causal inference from the multi-site intervention rather than merely establishing an association.^37–39^ Although the app was available within *Cerner* across all EDs beginning in April 2018, formal implementation, including structured advertisement via email and in-person or virtual training on how to use the app, did not begin until December 2019. RR, assisted by HH, led the formal implementation in partnership with the IU Health Chief Medical Information Officer (JS) and ED site directors. Implementation occurred in four waves, with each wave lasting two-months in duration and comprising three to four ED sites located in the same geographic region. Each ED served first as a control and subsequently as a test site, enabling causal inference about the effects of *Health Dart* on INPC use. *Health Dart* was implemented in four Wave 1 sites on December 13, 2019; three Wave 2 sites on February 7, 2020; three Wave 3 sites on June 8, 2020; and four Wave 4 sites on August 3, 2020. Implementation was paused for two months between Waves 2 and 3 due to the COVID-19 pandemic.

### Conceptual Models

We used the Unified Theory of Acceptance and Use of Technology (UTAUT) and Consolidated Framework for Implementation Research (CFIR) as our conceptual models. They identify implementation context (the setting and circumstances of the implementation) as one of the predominant indicators of user behaviors, adoption and sustainable use of HIE.^40,41,16^ In our analysis of the effect of the implementation of *Health Dart* on INPC use we focused on implementation context, which included enhancements to the UI, repositioning of the app in the toolbar, and the COVID-19 pandemic.

### Measures

We evaluated INPC use for the 286,175 ED encounters that occurred between October 1, 2019, and December 31, 2020, at all sites collectively. User log data and encounter details were extracted from the INPC and IU Health’s Enterprise Data Warehouse by the Regenstrief Institute Data Core. Encounters were categorized by implementation wave (Wave 1-4), INPC use (yes or no), and INPC access method (*CareWeb* or *Health Dart*). Encounters were also categorized as occurring pre-or post-implementation, before or after UI enhancements were made to the app (May 11, 2020), before or after the app was moved to a more prominent position on the toolbar (October 14, 2020), and before or during the COVID-19 pandemic as defined by the date of the first reported COVID-19 case in Indiana (March 6, 2020). To understand the temporal effects of app implementation, encounters were also grouped by three different time periods relative to implementation: pre-implementation, between 0 and 2 months post-implementation, and more than 2 months post-implementation. The outcome was a binary variable representing whether the INPC (either through *Health Dart* or *CareWeb*) was used in an encounter.

### Statistical Analysis

First, a descriptive analysis provided rates of INPC use at each ED site and implementation wave over the study period (October 1, 2019, to December 31, 2020). INPC use rates at each of the ED sites were calculated and plotted for each month of the study period to visualize the effects of *Health Dart* and implementation context (UI enhancement, toolbar repositioning, and COVID-19 pandemic) on INPC use. For each site, we also determined the proportion of INPC use that occurred through *Health Dart* versus *CareWeb*.

Second, we determined the associations between INPC use and variables describing the implementation context. Using a chi-square test, we compared INPC use rates between pre- and post-*Health Dart* implementation groups for all encounters and stratified them by the implementation wave, UI enhancements, toolbar repositioning, and COVID-19 pandemic. Chi-square and Cochran-Armitage Trend tests were used to compare and assess changes in INPC use rates across the three time periods (pre, 0-2 months, and >2 months) for all encounters, stratified by implementation wave, UI enhancements, and COVID-19 pandemic. A chi-square test was used to assess whether increases in INPC use were associated with the repositioning of the app on the toolbar. Because this change was made after *Health Dart* was implemented in all four waves, we did not compare INPC use rates among the three time periods while adjusting for the toolbar repositioning and could not separate the effect of the app implementation from the toolbar repositioning.

Third, logistic regression modeling was used to estimate the effect of *Health Dart* implementation on the probability of INPC use while adjusting for implementation wave, UI enhancements, and pandemic status. Specifically, we considered the following four models with different combinations of independent variables: Model A) the binary *Health Dart* implementation variable and implementation wave; Model B) variables in Model A, the binary variables of the UI enhancement, and pandemic status and their interactions with the *Health Dart* implementation variable; Model C) the three-level categorical variable of different *Health Dart* implementation time periods and implementation waves; and Model D) variables in Model C, the binary variables of the UI enhancement and pandemic status, and their interactions with the three-level categorical variable of different *Health Dart* implementation time periods. In all models, a site-level random intercept was included to account for clustering effects within the same sites.

A power analysis was conducted based on the stepped-wedge cluster non-randomized design and pilot study results to test the hypothesis that app implementation increased INPC use.^25^ A study site having at least 39 daily encounters per two-month implementation wave (a total of 2,340 encounters) would result in the conclusion that *Health Dart* led to greater INPC usage under a stringent test with a significance level of α ≤ 0.01 powered at 0.9. Given the large encounter volume at IU Health EDs (286,175 ED encounters) during the study period, there was adequate power to detect a statistically significant difference in INPC use due to the app implementation.

The implementation context is an important predictor of user behavior in the UTAUT and CFIR models, and we therefore modeled its influence on INPC use. The implementation context included introducing enhancements to the user interface (UI), repositioning the app to a more prominent location on the EHR toolbar, and the COVID-19 pandemic. The COVID-19 pandemic is an especially notable event as the National Syndromic Surveillance Program reported a 42% decline in ED visits between March 29 and April 25, 2020, compared to the same time period in 2019.^42^ During this time, many common ED chief complaints were displaced by infectious diseases and respiratory conditions compared to the prior year.^42,43^ In our study, we anticipated an incremental increase in overall INPC use at the beginning of the *Health Dart* implementation and a leveling off over time. We predicted a similar INPC use pattern following the UI enhancements and the repositioning of the app in the toolbar. Because there was a reduction in opportunities to use *Health Dart* for its intended purpose due to COVID-19, we expected an initial decrease in INPC use shortly after the start of the pandemic with a gradual increase and leveling off over time.

This study was reviewed and approved by the Indiana University Institutional Review Board (protocol #1905749709). Analyses were completed using SAS 9.4 (SAS Institute Inc., Cary, NC, USA). Statistical significance was determined using 95% confidence intervals (alpha level=0.05).

## RESULTS

### INPC Use by Implementation Wave

The average rate of INPC use at the 14 sites throughout the study period (October 1, 2019 to December 31, 2020) ranged from 1.2% in Wave 3 to 18.1% in Wave 1. Rates were similar among EDs in the same implementation wave except for IU Health Morgan in Wave 4, which used INPC at a higher rate than the other Wave 4 sites. In all waves, INPC use initially increased after implementation and continued to increase after UI enhancements and toolbar repositioning. The odds of INPC use increased 10% after the toolbar changes (p<0.001). In Wave 1, there was a decline in use of the INPC in March 2020 coinciding with the start of the COVID-19 pandemic with a gradual return to pre-pandemic rates over time; the effect of the pandemic was not as dramatic at the other sites (**Figure 2**). Across all four implementation waves, there was a pattern of incremental increases in INPC use in the 0 to 2 months and >2 months post-implementation time periods. At four sites (Tipton, Blackford, Jay, and Paoli), INPC use leveled off between the 0 to 2 months and >2 months post-implementation time periods (**Figure 3**).

**Figure 2.**
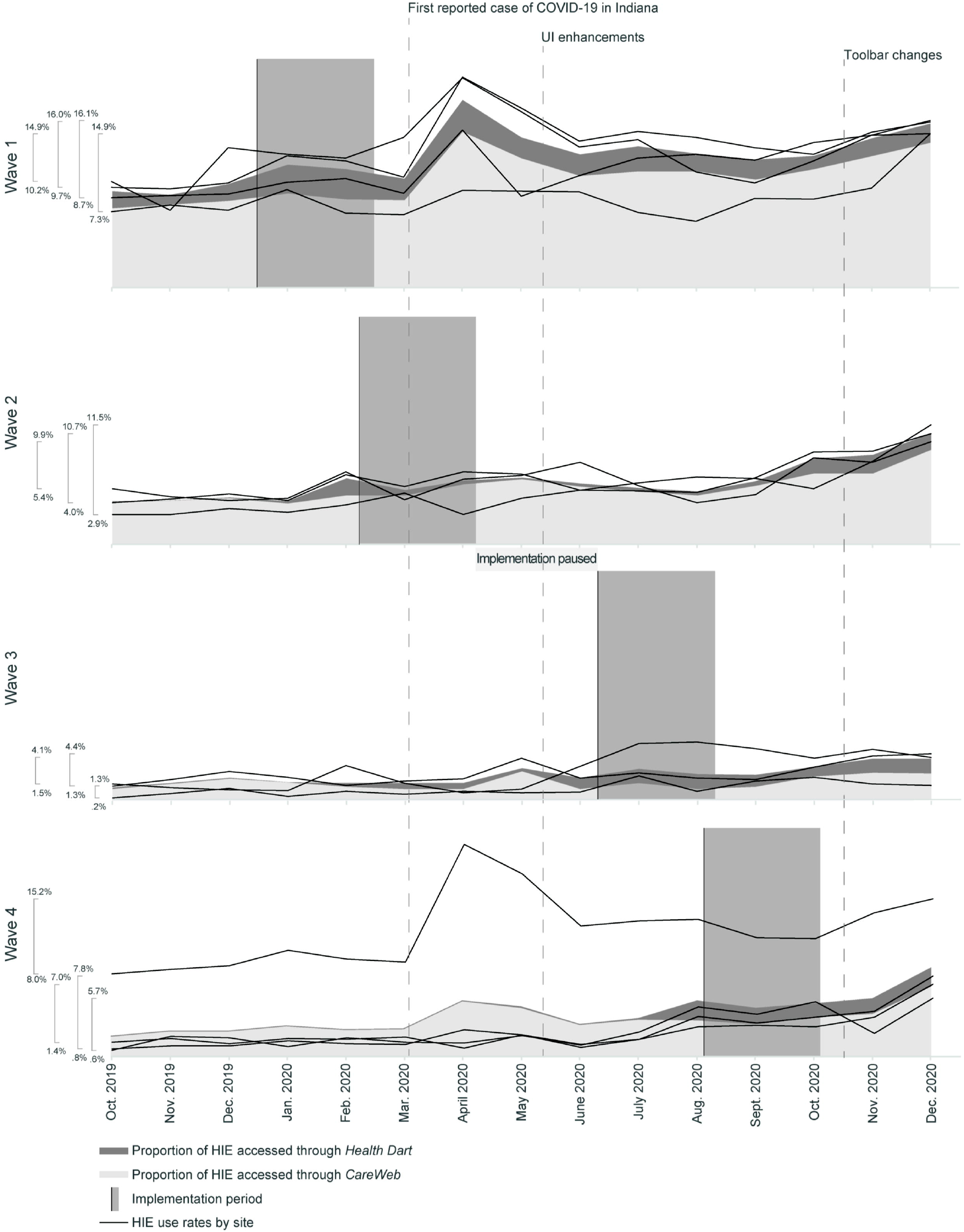
Monthly rates of HIE use in each *Health Dart* implementation wave. *Health Dart* was implemented in four Wave 1 sites on December 13, 2019; three Wave 2 sites on February 7, 2020; three Wave 3 sites on June 8, 2020; and four Wave 4 sites on August 3, 2020. The implementation study was temporarily paused between Wave 2 and Wave 3 due to the COVID-19 pandemic. 110 encounters had missing data (0.04%). The *Health Dart* application was available in *Cerner* in April 2018; therefore, we could not prevent use of the application prior to the study period for each wave. Abbreviations: UI, user interface; HIE, health information exchange.

**Figure 3.**
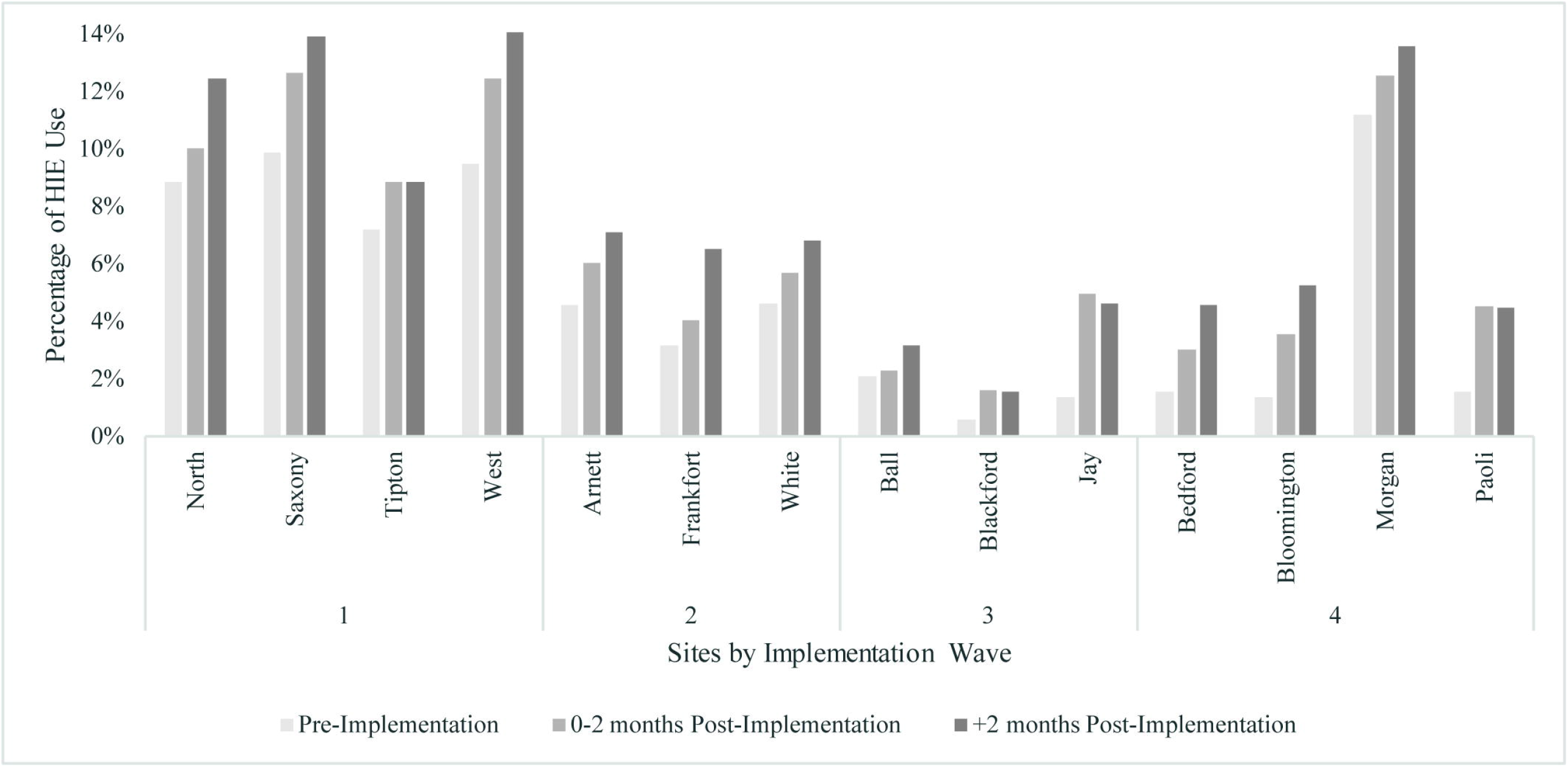
Rate of HIE use before and after *Health Dart* implementation at 14 IUH EDs in 4 implementation waves. HIE use combined access through the web-based portal and *Health Dart*.

### INPC Use Before and After *Health Dart* Implementation

Clinicians’ INPC use was 131% higher overall (from 3.6% to 8.3%; p<0.001) in post-implementation encounters compared to pre-implementation encounters (**Table 1**). The increased likelihood of INPC use in post-implementation encounters was also observed in each implementation wave, in pre- and post-UI enhancement periods, and before and during the COVID-19 pandemic. The effect of the implementation was not as strong in encounters that occurred after UI enhancements or in encounters that occurred during the pandemic. After UI enhancements, post-implementation INPC use increased 123% (from 3.5% to 7.8%; p<0.001) compared to 181% (from 3.6% to 10.1%; p<0.001) in post-implementation encounters that occurred before UI enhancements. During the pandemic, post-implementation INPC use increased by 135% (from 3.4% to 8.0%; p<0.001) compared to 178% (from 3.6% to 10%; p<0.001) in post-implementation encounters that occurred before the pandemic.

**Table 1.** Comparison of INPC use pre- and post-*Health Dart* implementation

| ED Encounters | N | (%) | Pre | Post | Increase | $\chi^2$ p-value | OR | [95% CI] |
| --- | --- | --- | --- | --- | --- | --- | --- | --- |
| Total | 286,175 | (100.0) | 3.6% | 8.3% | 131% | <0.001 | 2.44 | [2.35, 2.52] |
| Wave 1 | 72,639 | (25.4) | 9.2% | 13.0% | 41% | <0.001 | 1.48 | [1.39, 1.58] |
| Wave 2 | 63,667 | (22.2) | 4.4% | 6.7% | 52% | <0.001 | 1.58 | [1.46, 1.71] |
| Wave 3 | 61,715 | (21.6) | 1.8% | 3.0% | 67% | <0.001 | 1.73 | [1.56, 1.93] |
| Wave 4 | 88,154 | (30.8) | 3.1% | 5.9% | 90% | <0.001 | 1.95 | [1.82, 2.08] |
| Pre-UI enhancements | 143,024 | (50.0) | 3.6% | 10.1% | 181% | <0.001 | 3.03 | [2.89, 3.17] |
| Post-UI enhancements | 143,151 | (50.0) | 3.5% | 7.8% | 123% | <0.001 | 2.29 | [2.12, 2.49] |
| Pre-COVID-19 | 111,504 | (39.0) | 3.6% | 10.0% | 178% | <0.001 | 2.94 | [2.78, 3.12] |
| During COVID-19 | 174,671 | (61.0) | 3.4% | 8.0% | 135% | <0.001 | 2.50 | [2.35, 2.66] |

Data are derived from the Indiana University Health Data Warehouse covering 14 emergency departments. Total encounters were reported from October 1, 2019, to December 31, 2020. Encounters with missing data ( $n=110$ ; 0.04%) were excluded from analysis. Chi-square tests compared rates of INPC use between pre- and post-*Health Dart* implementation. Odds ratios represent the odds of INPC use post-*Health Dart* implementation divided by the odds of INPC use pre-*Health Dart* implementation.
*Note:* The overall percentage change may be very high even with relatively low percentage changes in the subgroups that have unbalanced numbers of encounters between pre-*Health Dart* and post-*Health Dart* implementation and varying group-specific INPC usage rates.
Abbreviations: CI, confidence interval; ED, emergency department; INPC, Indiana Network for Patient Care (health information exchange); OR, odds ratio; UI, user interface.

As was observed in the descriptive analysis, an INPC use rates over the pre-implementation, 0 to 2 months post-implementation, and >2 months post-implementation time periods increased across all encounters (p<0.001) (**Table 2**). This trend was also observed in each implementation wave, in pre- and post-UI enhancement periods, and before and during the COVID-19 pandemic. In each of the time periods, the effect of *Health Dart* on INPC use was not as strong in encounters that occurred after UI enhancements (p<0.001) or in encounters that occurred during COVID-19.

**Table 2.** Comparison of INPC use stratified by pre-, 0 to 2 months post, and more than 2 months post-*Health Dart* implementation time periods

| ED Encounters | N | (%) | Pre | *Health Dart* Time Periods | | $\chi^2$ p-value | CA p-value |
| --- | --- | --- | --- | --- | --- | --- | --- |
|  |  |  |  | 0-2 months post | >2 months post |  |  |
| Total | 286,175 | (100.0) | 3.6% | 6.5% | 8.8% | <0.001 | <0.001 |
| Wave 1 | 72,639 | (25.4) | 9.2% | 11.5% | 13.4% | <0.001 | <0.001 |
| Wave 2 | 63,667 | (22.2) | 4.4% | 5.6% | 6.9% | <0.001 | <0.001 |
| Wave 3 | 61,715 | (21.6) | 1.8% | 2.6% | 3.2% | <0.001 | <0.001 |
| Wave 4 | 88,154 | (30.8) | 3.1% | 5.0% | 6.4% | <0.001 | <0.001 |
| Pre-UI enhancements | 143,024 | (50.0) | 3.6% | 9.0% | 11.4% | <0.001 | <0.001 |
| Post-UI enhancements | 143,151 | (50.0) | 3.5% | 4.1% | 8.5% | <0.001 | <0.001 |
| Pre-COVID-19 | 111,504 | (39.0) | 3.6% | 9.9% | 10.3% | <0.001 | <0.001 |
| During COVID-19 | 174,671 | (61.0) | 3.4% | 4.3% | 8.8% | <0.001 | <0.001 |

Data were derived from the Indiana University Health Data Warehouse covering 14 emergency departments. Total encounters were reported from October 1, 2019, to December 31, 2020. Encounters with missing data ( $n = 110$ ; 0.04%) were excluded from analysis. To test for temporal trends in INPC use, three time periods were used: pre-*Health Dart* implementation, 0-2 months post-*Health Dart* implementation, and >2 months post-*Health Dart* implementation. Chi-square tests compared rates of INPC use between pre- and post-*Health Dart* implementation. Cochran-Armitage Trend tests determined whether INPC use increased over the three time periods.
Abbreviations: CA, Cochran-Armitage Trend test; ED, emergency department; INPC, Indiana Network for Patient Care (health information exchange); UI, user interface.

### Estimating the Probability of INPC Use

The odds of INPC use increased 68% post-*Health Dart* implementation, adjusted for the implementation wave (p<0.001) (**Table 3**). The odds of INPC use was 47% higher at 0 to 2 months post-implementation (p<0.001) and 76% higher at >2 months post-implementation (p<0.001) compared to pre-*Health Dart*, after controlling for wave.

**Table 3.** Modeling the probability of INPC use

| Model |  | OR | [95% CI] | p-value |
| --- | --- | --- | --- | --- |
| A | Post-*Health Dart* implementation | 1.68 | [1.62, 1.75] | <0.001 |
|  | Pre-*Health Dart* implementation | Reference |  |  |
| B | Post-*Health Dart* implementation at pre-UI enhancements | 1.38 | [1.29, 1.47] | <0.001 |
|  | Pre-*Health Dart* implementation at pre-UI enhancements | Reference |  |  |
|  | Post-*Health Dart* implementation at post-UI enhancements | 1.47 | [1.33, 1.64] | <0.001 |
|  | Pre-*Health Dart* implementation at post-UI enhancements | Reference |  |  |
|  | Post-*Health Dart* implementation at pre-COVID-19 | 1.46 | [1.33, 1.59] | <0.001 |
|  | Pre-*Health Dart* implementation at pre-COVID-19 | Reference |  |  |
|  | Post-*Health Dart* implementation during COVID-19 | 1.39 | [1.29, 1.50] | <0.001 |
|  | Pre-*Health Dart* implementation during COVID-19 | Reference |  |  |
| C | 0-2 months after *Health Dart* implementation | 1.47 | [1.40, 1.55] | <0.001 |
|  | >2 months after *Health Dart* implementation | 1.76 | [1.69, 1.83] | <0.001 |
|  | Pre-*Health Dart* implementation | Reference |  |  |
| D | 0-2 months after *Health Dart* implementation at pre-UI enhancements | 1.26 | [1.15, 1.39] | <0.001 |
|  | >2 months after *Health Dart* implementation at pre-UI enhancements | 1.37 | [1.26, 1.48] | <0.001 |
|  | Pre-*Health Dart* implementation at pre-UI enhancements | Reference |  |  |
|  | 0-2 months after *Health Dart* implementation at post-UI enhancements | 1.58 | [1.37, 1.81] | <0.001 |
|  | >2 months after *Health Dart* implementation at post-UI enhancements | 1.40 | [1.25, 1.57] | <0.001 |
|  | Pre-*Health Dart* implementation at post-UI enhancements | Reference |  |  |
|  | 0-2 months after *Health Dart* implementation at pre-COVID-19 | 1.64 | [1.46, 1.85] | <0.001 |
|  | >2 months after *Health Dart* implementation at pre-COVID-19 | 1.31 | [1.15, 1.48] | <0.001 |
|  | Pre-*Health Dart* implementation at pre-COVID-19 | Reference |  |  |
|  | 0-2 months after *Health Dart* implementation during COVID-19 | 1.21 | [1.09, 1.34] | 0.003 |
|  | >2 months after *Health Dart* implementation during COVID-19 | 1.46 | [1.35, 1.58] | <0.001 |
|  | Pre-*Health Dart* implementation during COVID-19 | Reference |  |  |

Abbreviations: ED, emergency department; INPC, Indiana Network for Patient Care (health information exchange); UI, user interface

The odds of INPC use was 38% higher after the *Health Dart* implementation and before UI enhancements (p<0.001). The effect of *Health Dart* strengthened after UI enhancement, with the odds increasing to 47% (p<0.001). There was an increasing trend of odds of INPC use over time post-*Health Dart* implementation both before and after UI enhancement. Before the UI enhancement, the odds of INPC use was 26% higher at 0 to 2 months post-implementation (p<0.001) and 37% higher at >2 months post-implementation (p<0.001) compared to pre-*Health Dart*. After the UI enhancement, the odds of INPC use was 58% higher at 0 to 2 months post-implementation and 40% higher at >2 months post-implementation (all p-values<0.001).

Before the COVID-19 pandemic, the odds of INPC use was 46% higher after *Health Dart* implementation (p<0.001). The effect of *Health Dart* slightly weakened during the COVID-19 pandemic, when the odds of INPC use was 39% higher after *Health Dart* implementation (p<0.001). Before the pandemic, the odds INPC use was 64% higher at 0 to 2 months post-implementation and 31% higher at >2 months post-implementation compared to pre-implementation (all p-values <0.001). On the other hand, during the pandemic, there was a clear trend of increasing *Health Dart* effect over time, with the odds of INPC use 21% higher at 0 to 2 months post-implementation (p=0.003) and 46% higher at >2 months post-implementation (p<0.001).

## DISCUSSION

This study had two major goals: (1) develop *Health Dart*, a FHIR app that integrates information from the INPC directly with *Cerner* in the form of a chief complaint-oriented dashboard; (2) determine how the implementation of *Health Dart* affected INPC use among clinicians at 14 IU Health EDs using a stepped-wedge trial design. The development of *Health Dart* addressed two current, major limitations of HIE implementation and use. First, it integrated relevant information from the HIE directly into the EHR as opposed to forcing clinicians to access a separate HIE system. Second, it presented this information in the form of a dashboard focused on a chief complaint, obviating the need for the clinician to manually collate this information.

As a result of the implementation of *Health Dart*, INPC use increased by 131% (from 3.6% to 8.3%; p<0.001) overall and by 144% (from 3.6% to 8.8%; p<0.001) more than 2 months after implementation. This increase in INPC use in one health system was remarkable considering the 9% increase for EDs across multiple Indiana health systems reported during the period between 2011 and 2017 subsequent to the passage of the 2009 HITECH Act and UI enhancements.^44^ Our results support the benefits of direct integration of HIE information into an EHR for HIE use. This is consistent with previous findings that aggregation of data from disparate sources into a single view increased the frequency with which ED clinicians accessed HIE by 91.7%.^26^ In addition, the display of data in a structured, problem-oriented format, such as *Health Dart*’s chief complaint view, improves efficiency of data retrieval and user satisfaction.^28,45,46^

Contrary to our prediction, the effect of the app’s implementation on INPC use was not as strong in encounters that occurred after UI enhancements. The UI enhancements appeared to have had a temporal effect on INPC use, with an initial increase in INPC use in the short term (<2 months) followed by a return to pre-UI enhancement INPC use rates in the medium term (>2 months). The gradual effect of *Health Dart* implementation and UI enhancements on INPC use is supported by the Innovation Diffusion Theory, in which the use of technology increases slowly at first and then rapidly before leveling off.^47^ The individual, social, and organizational factors outlined in the UTAUT model also can potentially explain why INPC use rates and methods of use differed by ED and by implementation wave. That is, clinicians in each of the EDs have different expectations (e.g. leaders, peers, and organizational and technical infrastructures) that influence their intention and behavior to use *Health Dart*.^40^ Variations in rates of INPC use across sites and implementation waves is also consistent with EHR implementation evaluations which note how variability in implementation processes, such as physician training and timing of software updates, influence clinicians’ perceptions about the usability of a newly implemented technology.^48^ In our study, which focused on implementation context guided by the CFIR model, the implementation factors (i.e., UI enhancements and COVID-19 pandemic) weakened the effect of *Health Dart* in increasing INPC use.

Moving the *Health Dart* app toward the top of the *Cerner PowerChart* toolbar was associated with an increased likelihood of INPC use among ED clinicians. Relocation of the app to a more prominent location presumably increased its usability by decreasing the time required for users to find and use it. This finding is consistent with Fitt’s law and Jakob Nielsen’s usability heuristics for user interface design. Fitt’s law describes the amount of time it takes a user to complete an action as a function of the distance and accuracy of the movement, while Nielsen’s usability heuristics suggest that visibility and prioritization of relevant content increase usability.^49,50^ Because the toolbar relocation occurred toward the end of the study, we were not able to separate the effects of the *Health Dart* implementation on INPC use from the effects of the toolbar changes. To reduce confounding of our results, we excluded encounters that occurred prior to *Health Dart* implementation when evaluating the effect of toolbar relocation on INPC use.

Congruent with our prediction, the effect of the implementation on INPC use was not as strong in encounters that occurred during as opposed to prior to the COVID-19 pandemic. The pandemic had a temporal effect on INPC use, with a decline in INPC use in the short term (<2 months) followed by a return to pre-COVID-19 INPC use rates in the medium term (>2 months). The results of the COVID-19 modeling should be interpreted with caution as the lower volume of patients and higher rate of infectious diseases and respiratory conditions in encounters during the pandemic increase the potential for confounding.

This study extends our prior work through a larger sample size, increased statistical power, and the ability to limit the effect of confounding using the stepped-wedge trial design.^25,44^ Although the data were derived from only one statewide hospital system in the Midwest, this novel method can potentially be scaled to additional EDs and HIEs to improve generalizability of results for different populations, hospital systems, and EHR vendor platforms. The comprehensive and diverse data sources of the INPC also serve to improve the generalizability of our results. The INPC connects 123 hospitals from 38 health systems, 19,095 clinical practices and 54,107 providers, and represents more than 19 million patients and 16 billion clinical data elements.^51^ In current research, we plan to analyze the probability of INPC use given clinician, patient, and encounter characteristics; the utility of INPC access given various encounter, patient and clinician characteristics; and how clinicians decide to access *Health Dart, CareWeb* or both. A more granular level of analysis may elucidate anomalies, such as IU Health Morgan’s higher rates of INPC use compared to other Wave 4 sites. Additionally, because *Health Dart* was designed for seven chief complaints, we will examine whether there is a relationship between chief complaint and use of INPC. The user log data indicate if a user accessed INPC data, but to date we are unable to account for what information the user accessed and whether it was relevant to clinical decision-making, care provided or clinical outcomes. Knowing what the user accessed could inform whether the actual rates of INPC use reflect the opportunities for appropriate use. Currently, how to determine optimal levels of HIE use in the ED remains unknown.

### Limitations

Similar to other evaluations of healthcare technology adoption and use, this study did not measure the variability among individual clinicians when assessing the causal effect of the *Health Dart* implementation on INPC use.^18-20^ Models controlled for ED sites to account for differences between individual clinicians, and we assumed that clinicians working at the same ED had similar organizational and social influences.

## CONCLUSION

Our results provide support for the benefits of directly integrating HIE information into the EHR in a problem-oriented format for promoting HIE use in the ED. In addition, they provide evidence of the influence of implementation context in the adoption and use of HIE. The results underscore the importance of considering contextual influences such as culture, policy, and setting when evaluating the implementation of a novel technology in the ED.

## Data Availability

All data produced in the present study are available upon reasonable request to the authors

## ACKNOWLEDGEMENTS

The authors acknowledge Drs. Richard Holden, Joshua Vest, Julia Adler-Milstein, and Saurabh Rahurkar for their contributions to this research. Thank you to Sarah Zappone, Laura Ruppert, Abena Gyasiwa, and Emily Fortier for their coordination of this work. The authors also thank IU Health, the IU Health ED clinicians, and Indiana Health Information Exchange for their participation in the study.

## Footnotes

1 \**CareWeb* uses single sign-on through *Cerner*, typically obviating the need for clinicians to log in separately.

## REFERENCES

1. Adler-Milstein J, Embi PJ, Middleton B, et al. Crossing the health IT chasm: Considerations and policy recommendations to overcome current challenges and enable value-based care. J Am Med Inform Assoc. 2017;24(5):1036–1043. doi:10.1093/jamia/ocx017

2. Lichtner V, Baysari M. Electronic display of a patient treatment over time: A perspective on clinicians’ burn-out. BMJ Health Care Inform. 2021;28(1):e100281. doi:10.1136/bmjhci-2020-100281

3. Friedberg MW, Chen PG, Van Busum KR, et al. Factors affecting physician professional satisfaction and their implications for patient care, health systems, and health policy. Rand Health Q. 2014;3(4):1. http://www.ncbi.nlm.nih.gov/pubmed/28083306. Accessed March 15, 2022.

4. Payne TH, Corley S, Cullen TA, et al. Report of the AMIA EHR-2020 Task Force on the status and future direction of EHRs. J Am Med Inform Assoc. 2015;22(5):1102–1110. doi:10.1093/jamia/ocv066

5. Bourgeois FC, Olson KL, Mandl KD. Patients treated at multiple acute health care facilities: Quantifying information fragmentation. Arch Intern Med. 2010;170(22):1989–1995. doi:10.1001/archinternmed.2010.439

6. Windle JR, Katz AS, Dow JP, et al. 2016 ACC/ASE/ASNC/HRS/SCAI health policy statement on integrating the healthcare enterprise. J Am Coll Cardiol. 2016;68(12):1348–1364. doi:10.1016/j.jacc.2016.04.017

7. Menachemi N, Rahurkar S, Harle CA, et al. The benefits of health information exchange: An updated systematic review. J Am Med Inform Assoc. 2018;25(9):1–7. doi:10.1093/jamia/ocy035

8. Rudin RS, Motala A, Goldzweig CL, et al. Usage and effect of health information exchange: A systematic review. Ann Intern Med. 2014;161(11):803–811. doi:10.7326/M14-0877

9. Patel V, Henry J, Pylypchuk Y, et al. Interoperability among U.S non-federal acute care hospitals in 2015. ONC Data Brief. 2016; 36(1):1–11. https://www.healthit.gov/sites/default/files/briefs/onc_data_brief_36_interoperability.pdf. Accessed March 15, 2022.

10. The Office of the National Coordinator for Health Information Technology. Final interoperability roadmap statements of support. http://HealthIT.gov. https://www.healthit.gov/sites/default/files/interoperability_roadmap_statements_of_support_2015-11-16_1.pdf. Accessed Martch 15, 2022.

11. Centers for Medicare and Medicaid Services. MACRA: MIPS & APMs. http://CMS.gov https://www.cms.gov/medicare/quality-initiatives-patient-assessment-instruments/value-based-programs/macra-mips-and-apms/macra-mips-and-apms. Accessed March 15, 2022.

12. Holmgren AJ, Adler-Milstein J. Health Information Exchange in U.S. hospitals: The current landscape and a path to improved information sharing. J Hosp Med. 2017;12(03):193–198. doi:10.12788/jhm.2704

13. Upton F. 21st Century Cures Act, H.R. 6. 114^th^ Cong. (2015-2016). https://www.congress.gov/bill/114th-congress/house-bill/6/text. Accessed March 15, 2022.

14. Pronovost P, Palmer S, Johns, MME, et al. Procuring interoperability: Achieving high-quality, connected, and person-centered care. National Academy of Medicine. https://nam.edu/procuring-interoperability-achieving-high-quality-connected-and-person-centered-care/. Accessed March 15, 2022.

15. Ruley M, Walker V, Studeny J, et al. The nationwide health information network: The case of the expansion of health information exchanges in the United States. Health Care Manag (Frederick). 2018;37(4):333–338. doi:10.1097/HCM.0000000000000231

16. Lin SC, Everson J, Adler-Milstein J. Technology, incentives, or both? Factors related to level of hospital health information exchange. Health Serv Res. 2018;53(5):3285–3308. doi:10.1111/1475-6773.12838

17. Kibbe DC, Phillips RL, Green LA. The Continuity of Care Record. Am Fam Physician. 2004;70(7):1220–1223. https://www.aafp.org/afp/2004/1001/p1220.html. Accessed March 15, 2022.

18. Stack SJ, Botstein G, Mattison J, et al. Improving care: Priorities to improve electronic health record usability. American Medical Association. https://www.ama-assn.org/sites/ama-assn.org/files/corp/media-browser/member/about-ama/ehr-priorities.pdf. Accessed March 15, 2022.

19. Smith PC, Araya-Guerra R, Bublitz C, et al. Missing clinical information during primary care visits. JAMA. 2005;293:565–571. doi:10.1001/jama.293.5.565

20. Everson J. The implications and impact of 3 approaches to health information exchange: Community, enterprise, and vendor-mediated health information exchange. Learn Health Syst. 2017;1(2):1–9. doi:10.1002/lrh2.10021

21. American Hospital Association. Sharing health information for treatment. TrendWatch. https://www.aha.org/system/files/2018-03/sharing-health-information.pdf. Accessed March 15, 2022.

22. Furukawa MF, King J, Patel V, et al. Despite substantial progress in EHR adoption, health information exchange and patient engagement remain low in office settings. Health Aff (Millwood). 2014;33(9):1672–1679. doi:10.1377/hlthaff.2014.0445

23. Holmgren AJ, Patel V, Adler-Milstein J. Progress in interoperability: Measuring US hospitals’ engagement in sharing patient data. Health Aff (Millwood). 2017;36(10):1820–1827. doi:10.1377/hlthaff.2017.0546

24. Jensen LG, Bossen C. Factors affecting physicians’ use of a dedicated overview interface in an electronic health record: The importance of standard information and standard documentation. Int J Med Inform. 2016;87:44–53. doi:10.1016/j.ijmedinf.2015.12.009

25. Schleyer TKL, Rahurkar S, Baublet AM, et al. Preliminary evaluation of the Chest Pain Dashboard, a FHIR-based approach for integrating health information exchange information directly into the clinical workflow. AMIA Jt Summits Transl Sci Proc. 2019;2019:656–664. https://www.ncbi.nlm.nih.gov/pmc/articles/PMC6568135/. Accessed March 15, 2022.

26. Adler-Milstein J, Wang MD. The impact of transitioning from availability of outside records within electronic health records to integration of local and outside records within electronic health records. J Am Med Inform Assoc. 2020;27(4):606–612. doi:https://doi.org/10.1093/jamia/ocaa006

27. Buchanan J. Accelerating the benefits of the problem oriented medical record. Appl Clin Inform. 2017;8(1):180–190. doi:10.4338/ACI-2016-04-IE-0054

28. Semanik MG, Kleinschmidt PC, Wright A, et al. Impact of a problem-oriented view on clinical data retrieval. J Am Med Inform Assoc. 2021;28(5):899–906. doi:10.1093/jamia/ocaa332

29. Koopman RJ, Kochendorfer KM, Moore JL, et al. A diabetes dashboard and physician efficiency and accuracy in accessing data needed for high-quality diabetes care. Ann Fam Med. 2011;9(5):398–405. doi:10.1370/afm.1286

30. Kummer BR, Willey JZ, Zelenetz MJ, et al. Neurological dashboards and consultation turnaround time at an academic medical center. Appl Clin Inform. 2019;10(5):849–858. doi:10.1055/s-0039-1698465

31. Khairat SS, Dukkipati A, Lauria HC, et al. The impact of visualization dashboards on quality of care and clinician satisfaction: Itegrative literature review. JMIR Hum Factors. 2018;5(2):e22. doi:10.2196/humanfactors.9328

32. Yan Q, Jiang Z, Harbin Z, et al. Exploring the relationship between electronic health records and provider burnout: A systematic review. J Am Med Inform Assoc. 2021;28(5):1009–1021. https://academic.oup.com/jamia/article/28/5/1009/6154416. Accessed March 15, 2022.

33. Cantwell E, Mcdermott K. Making technology talk: How interoperability can improve care, drive efficiency, and reduced waste. Healthc Financ Manage. 2016;70(5):70–76. https://pubmed.ncbi.nlm.nih.gov/27382711/. Accessed March 15, 2022.

34. Ely JW, Osheroff JA, Chambliss ML, et al. Answering physicians’ clinical questions: Obstacles and potential solutions. J Am Med Inform Assoc. 2005;12(2):217–224. doi:10.1197/jamia.M1608

35. Indiana Health Information Exchange. OneCare. https://www.ihie.org/onecare/#careweb. Accessed March 15, 2022.

36. Rui P, Kang K. National Hospital Ambulatory Medical Care Survey: 2017 emergency department summary tables. National Center for Health Statistics. https://www.cdc.gov/nchs/data/nhamcs/web_tables/2017_ed_web_tables-508.pdf. Accessed March 15, 2022.

37. Barker D, McElduff P, D’Este C, et al. Stepped wedge cluster randomised trials: A review of the statistical methodology used and available. BMC Med Res Methodol. 2016;16(1):1–19. doi:10.1186/s12874-016-0176-5

38. Hussey MA, Hughes JP. Design and analysis of stepped wedge cluster randomized trials. Contemp Clin Trials. 2007;28(2):182–191. doi:10.1016/j.cct.2006.05.007

39. Hemming K, Haines TP, Chilton PJ, et al. The stepped wedge cluster randomised trial: Rationale, design, analysis, and reporting. BMJ. 2015;350(1):391. doi:10.1136/bmj.h391

40. Venkatesh V, Morris M, Davis G, et al. User acceptance of information technology: Toward a unified view. MIS Q. 2003;27(3):425. doi:10.2307/30036540

41. Damschroder LJ, Aron DC, Keith RE, et al. Fostering implementation of health services research findings into practice: A consolidated framework for advancing implementation science. Implement Sci. 2009;4(1). doi:10.1186/1748-5908-4-50

42. Hartnett KP, Kite-Powell A, Devies J, et al. Impact of the COVID-19 Pandemic on emergency department visits — United States, January 1, 2019–May 30, 2020. MMWR Morb Mortal Wkly Rep. 2020;69(23):699–704. doi:http://dx.doi.org/10.15585/mmwr.mm6923e1

43. Venkatesh AK, Janke AT, Shu-Xia L, et al. Emergency department utilization for emergency conditions during COVID-19. Ann Emerg Med. 2021;78(1):84–91. doi:10.1016/j.annemergmed.2021.01.011

44. Rahurkar S, Vest JR, Finnell JT, Dixon BE. Trends in user-initiated health information exchange in the inpatient, outpatient, and emergency settings. J Am Med Informatics Assoc. 2021;28(3):622–627. doi:10.1093/jamia/ocaa226

45. Thayer JG, Ferro DF, Miller JM, et al. Human-centered development of an electronic health record-embedded, interactive information visualization in the emergency department using fast healthcare interoperability resources. J Am Med Inform Assoc. 2021;28(7):1401–1410. doi:https://.doi.org/10.1093/jamia/ocab016

46. Curran RL, Kukhareva PV, Taft T, et al. Integrated displays to improve chronic disease management in ambulatory care: A SMART on FHIR application informed by mixed-methods user testing. J Am Med Inform Assoc. 2020;27(8):1225–1234. doi:https://.doi.org/10.1093/jamia/ocaa099

47. Rogers EM. Diffusion of Innovations. 3<sup>rd</sup> ed. New York, NY: Free Press; 1983.

48. Ratwani RM, Savage E, Will A, et al. A usability and safety analysis of electronic health records: A multi-center study. J Am Med Inform Assoc. 2018;25(9):1197–1201. doi:10.1093/jamia/ocy088

49. Nielsen J. Usability Engineering. San Francisco, CA: Morgan Kaufmann Publishers Inc.; 1994.

50. Fitts PM. The information capacity of the human motor system in controlling the amplitude of movement. J Exp Psychol. 1954;47(6):381–391. http://www2.psychology.uiowa.edu/faculty/mordkoff/InfoProc/pdfs/Fitts1954.pdf. Accessed March 15, 2022.

51. Indiana Health Information Exchange. http://IHIE.org. https://www.ihie.org/. Accessed March 15, 2022.

